# NutriConnect: Enhancing Health and Food Security through Sustainable Solutions and Partnerships: Design and Protocol of a Pragmatic Comparative Effectiveness Trial

**DOI:** 10.1101/2025.05.20.25327991

**Authors:** Sang Gune K. Yoo, Rachel G. Tabak, Stephanie Mazzucca-Ragan, Allison Primo, Doneisha Bohannon, Derek Hashimoto, Charles W. Goss, Jason Wu, Eghonghon Eromosele, Ihab Hassanieh, Adam Hively, Abygail Martinez, Jinli Wang, Mark D. Huffman, Jing Li

## Abstract

**Background:** Food insecurity and poor dietary intake contribute to health disparities, particularly among socioeconomically disadvantaged populations. Produce prescription programs aim to improve access to fruits and vegetables (F&V) for those with diet-sensitive conditions, but comparative effectiveness data are limited.

**Objective:** To compare the impact of two produce prescription strategies, NutriConnect Credit (grocery coupons) and NutriConnect Delivery (home-delivered F&V boxes), on dietary intake, food security, and health outcomes among socioeconomically disadvantaged populations who have been recently hospitalized with diet-sensitive conditions.

**Methods:** In this three-arm pilot trial, recently hospitalized adults with food or financial insecurity and elevated cardiovascular risk were randomized (1:1:1) to Credit, Delivery, or enhanced usual care. The primary outcome is between group difference in change in F&V intake at 6 months. Secondary outcomes include food security and self-reported health-related quality of life. Implementation outcomes are assessed using the PRISM/RE-AIM framework.

**Conclusion:** NutriConnect seeks to provide evidence on the effectiveness and feasibility of two produce prescription strategies to inform scalable “Food is Medicine” programs targeting nutrition-related health disparities.

**Trial registration number:** NCT06263751

## Introduction

Food insecurity is a critical public health issue closely tied to poor dietary intake and diet-related chronic diseases, disproportionately affecting socioeconomically disadvantaged populations, including racial and ethnic minorities.^1–4^ To address food insecurity and diet-related chronic diseases, produce prescription programs, a Food is Medicine (FIM) strategy, aim to improve health outcomes by increasing access to fruits and vegetables (F&V) for individuals with limited resources and diet-sensitive health conditions.^5^ Produce prescriptions are defined as vouchers or discounted produce and groceries made available to people with the intention of increasing their consumption of healthier foods and improving their health.^6^ Systematic reviews highlight the potential of these programs to increase F&V intake, reduce food insecurity, and improve diet-related health conditions.^7,8^ However, variations in program design and inconsistent effect sizes of Food is Medicine interventions on health outcomes limits the reliability of the current evidence base.^9,10^

Despite growing interest and research related to produce prescription programs, the limited number of robust comparative trials also makes it challenging to determine the most effective strategies for patients who are socioeconomically disadvantaged.^12^ Additionally, many healthcare systems lack the infrastructure and experience to scale these initiatives effectively.^9,10^ Additionally, barriers such as cost, accessibility, and logistical complexities continue to hinder equitable distribution.^10,11^ The NutriConnect trial seeks to address these limitations by evaluating two approaches: (1) NutriConnect Credit, which provides participants with biweekly grocery coupons for purchasing F&V, and (2) NutriConnect Delivery, which offers preselected F&V boxes delivered biweekly to participants’ homes, both compared with usual care.^13^ We sought to assess the effects of both NutriConnect Credit and NutriConnect Delivery on F&V intake, food security, and self-reported health status. Additionally, we sought to evaluate the feasibility, reach, and fidelity in St. Louis metropolitan area where one in seven adults need food assistance, disproportionately affecting the Black population.^14,15^

Herein, we present the design, protocol, and baseline characteristics for a three-arm randomized (1:1:1) trial evaluating the comparative effectiveness of two produce prescription strategies, compared with enhanced usual care, in improving F&V intake and reducing food insecurity at six months among socioeconomically disadvantaged populations who have been recently hospitalized with diet-sensitive conditions.

### Study Design

The NutriConnect study is a comparative effectiveness trial designed as part of the rapid-cycle trials initiative supported by the American Heart Association Health Care by Food Program, aiming to generate insights that will inform future outcomes-driven trials of Food is Medicine interventions.^5^ The study consisted of a 13-month recruitment period followed by a 6-month intervention phase. NutriConnect seeks to address this gap by conducting a three-arm, randomized (1:1:1) comparative effectiveness study that compares two produce prescription strategies with enhanced usual care. In addition to assessing the interventions’ impact on F&V intake and household food security, the study explores key implementation outcomes such as reach, fidelity, implementation cost, and maintenance of these produce prescriptions strategies. Furthermore, this study compares the separate and combined effects of these strategies in populations most affected by food insecurity, particularly individuals facing transportation barriers or limited access to fresh produce. The study was approved by the institutional review board at Washington University in St. Louis (#202401096) and was prospectively registered (NCT06263751).

### Setting

The target population for this study is patients discharged from a large, academic hospital in St. Louis, Missouri. Barnes Jewish Hospital is a 1400-bed patient care, teaching facility and cares for a large and diverse patient population. Over one-fourth (27%) of patients self-identified as racial or ethnic minorities. In partnership with Schnucks, a regional grocery store with 200+ establishments serving over 20 million US residents, NutriConnect provides two different types of produce prescriptions further described below. The St. Louis metropolitan area faces persistent challenges with food access and insecurity, compounded by significant racial disparities both locally and across Missouri.^14,15^ NutriConnect aims to promote community health and well-being and address blatant disparities in our community.

### Implementation Strategies

NutriConnect is a collaborative consortium that includes Washington University in St. Louis, BJC HealthCare, and Schnucks. We previously developed and tested this collaboration through a pilot project, called the HealthyLink Model, which provided a three-day supply of food to patients experiencing food insecurity upon hospital discharge. Using heuristic evaluation, cognitive walkthroughs, and co-creation sessions with stakeholders, HealthyLink prioritized patient choice and tailored the intervention arms to accommodate participants’ health, cultural, and religious preferences. Incorporating inputs from partners and insights gained from the HealthyLink model, NutriConnect strategies were built upon the lived experience of target population and hospital frontline practice process, representing the next step in designing, implementing, and evaluating a Food is Medicine program in St. Louis.

NutriConnect interventions include two types of produce prescription strategies: NutriConnect Credit and NutriConnect Delivery. Patients randomized to the NutriConnect Credit arm received a $20 digital coupon every other week for purchasing F&V. This coupon was delivered via email and was restricted to food items recommended by a dietician (AP). A coupon was chosen instead of a digital reward applied directly to a store account because digital credit would not have restricted purchases, potentially allowing spending on unhealthy items. Participants randomized to the NutriConnect Delivery arm received a produce (F&V) box, equivalent in value to $20, delivered to their home every other week. The contents of the box were pre-selected through a collective effort of a Schnucks dietician (AP), the NutriConnect research team, and community partners to ensure a diverse and nutritious assortment of produce was made available. We selected the $20 biweekly allocation, equivalent to approximately $43 per month, based on a comprehensive review of six studies focused on adults, which reported monthly allocations ranging from $15 to $90 adults.^13^ The delivery was completed by Instacart, an online grocery delivery service and Schnucks partner. Due to the rising cost of produce and to account for participant convenience and seasonality of produce, for both NutriConnect Credit and Delivery groups, canned and frozen F&V were made available as deemed healthy by the study dietician (AP).

### Participants

Participants in the NutriConnect trial needed to meet specific eligibility criteria to ensure the intervention targeted the intended population. Eligible participants were adults (≥18 years) who received care on medical floors or in the observation unit at Barnes Jewish Hospital and were discharged to home. To be eligible for the study, participants had to screen positive for food or financial insecurity using either a validated two-item food insecurity screening questionnaire or by responding ‘very hard,’ ‘hard,’ or ‘somewhat hard’ to the question, “How hard is it for you to pay for the very basics like food, housing, heating, medical care, and medications?”^16^ Participants also had to have a history of elevated cardiovascular risk, defined as self-reported history of diabetes, hypertension, hyperlipidemia, or a body mass index of 30 kg/m^2^ or greater (Figure 1). Finally, participants were required to reside within the Schnucks delivery area. They were excluded from the study if they lacked the ability to provide informed consent, were under suicide watch or in police custody, were receiving hospice or palliative care, did not have a stable home, were pregnant, or had a chronic condition requiring a special diet (e.g., end-stage renal disease). As part of the routine discharge process, social workers asked for patients’ verbal permission to be contacted by trained members of the NutriConnect study team. The study team worked closely with the social workers to call patients post-discharge within 30 days. Trained study staff introduced the study, explained the enrollment process, and provided details on the risks and requirements of participation. If the patient agreed to participate, then study staff obtained and documented verbal consent prior to randomization.

**Figure 1.**
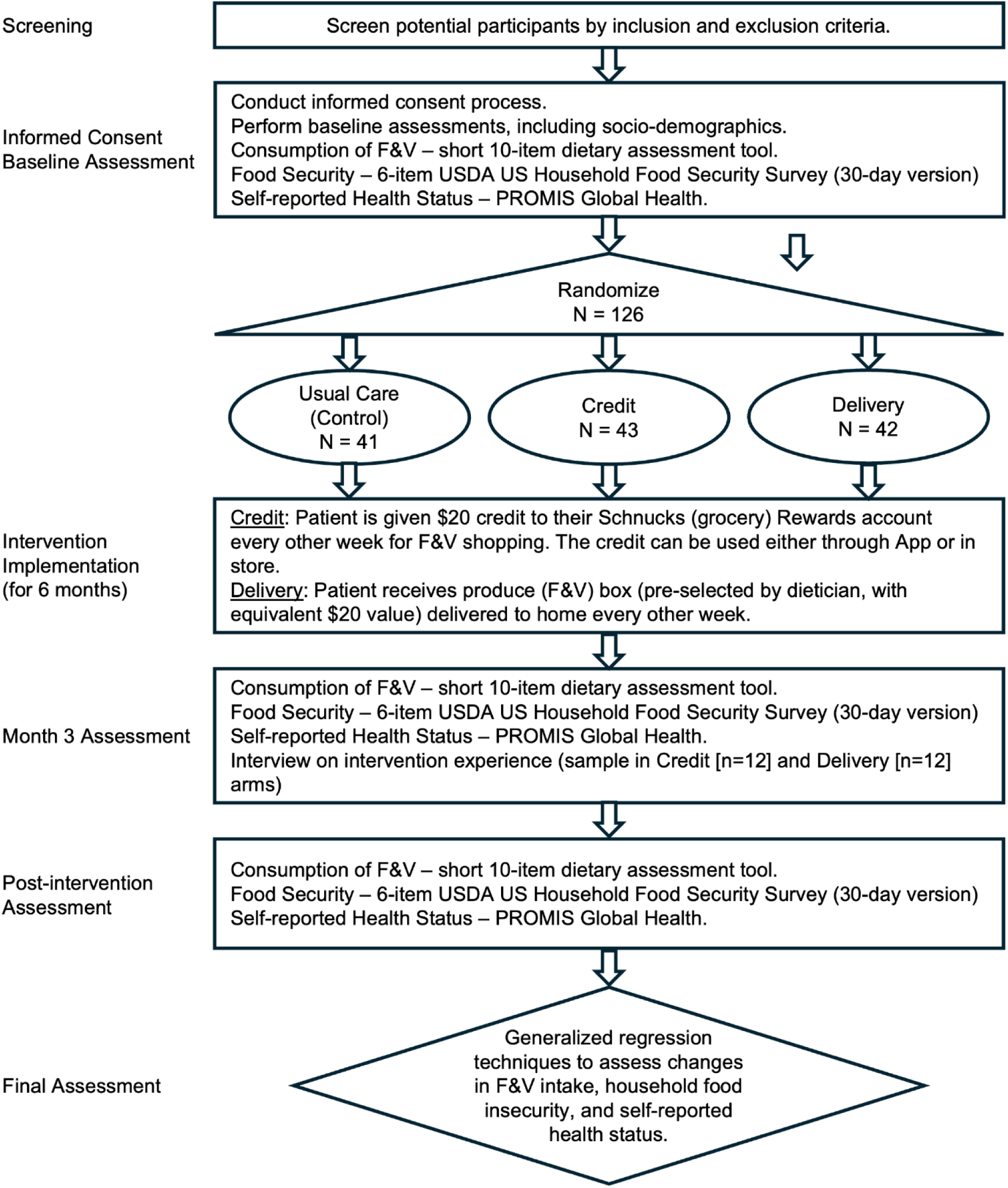
Trial design schematic.

All study participants needed to have or to sign up for a Schnucks Reward account. Since the registration process required minimal information, research staff and Schnucks customer care team assisted patients in setting up an account after the enrollment if they did not already have one. For patients randomized to the NutriConnect Credit, the $20 digital coupon could be redeemed for a single in-store, online, or Schnucks’ mobile app purchase. Participants randomized to the NutriConnect Delivery arm needed to have the ability to receive a produce box at home. To facilitate deliveries, the NutriConnect team confirmed the participant’s home address and preferred delivery day and time and collected two phone numbers from the participant along with two additional contacts for delivery notifications. Every other week, participants received text messages to inform them of the delivery window and status updates. Participants randomized to the enhanced usual care group receive the standard hospital-based approach to addressing food and financial insecurity, referring them to existing community service programs and food pantries. This arm serves as a control to evaluate the added value of structured Food Is Medicine interventions beyond routine social care referrals.

Randomization was implemented via a stratified random allocation scheme uploaded into REDCap. The stratification factor was race/ethnicity to ensure balance across the three arms. Additionally, permuted block randomization was implemented using variable block sizes to maintain temporal balance in arm allocation. The random allocation table was generated prior to study initiation by the study biostatistician (CWG) and stored in a password-protected REDCap project.

### Outcomes

Outcome assessments are planned at baseline, 3 months, and 6 months post-randomization (Table 1). To promote retention and encourage timely completion of follow-up assessments, NutriConnect provides an incremental incentive structure. Participants will receive a $10 gift card following completion of the baseline survey, a $15 gift card after the 3-month survey, and a $20 gift card upon completing the 6-month follow-up survey.

**Table 1.**
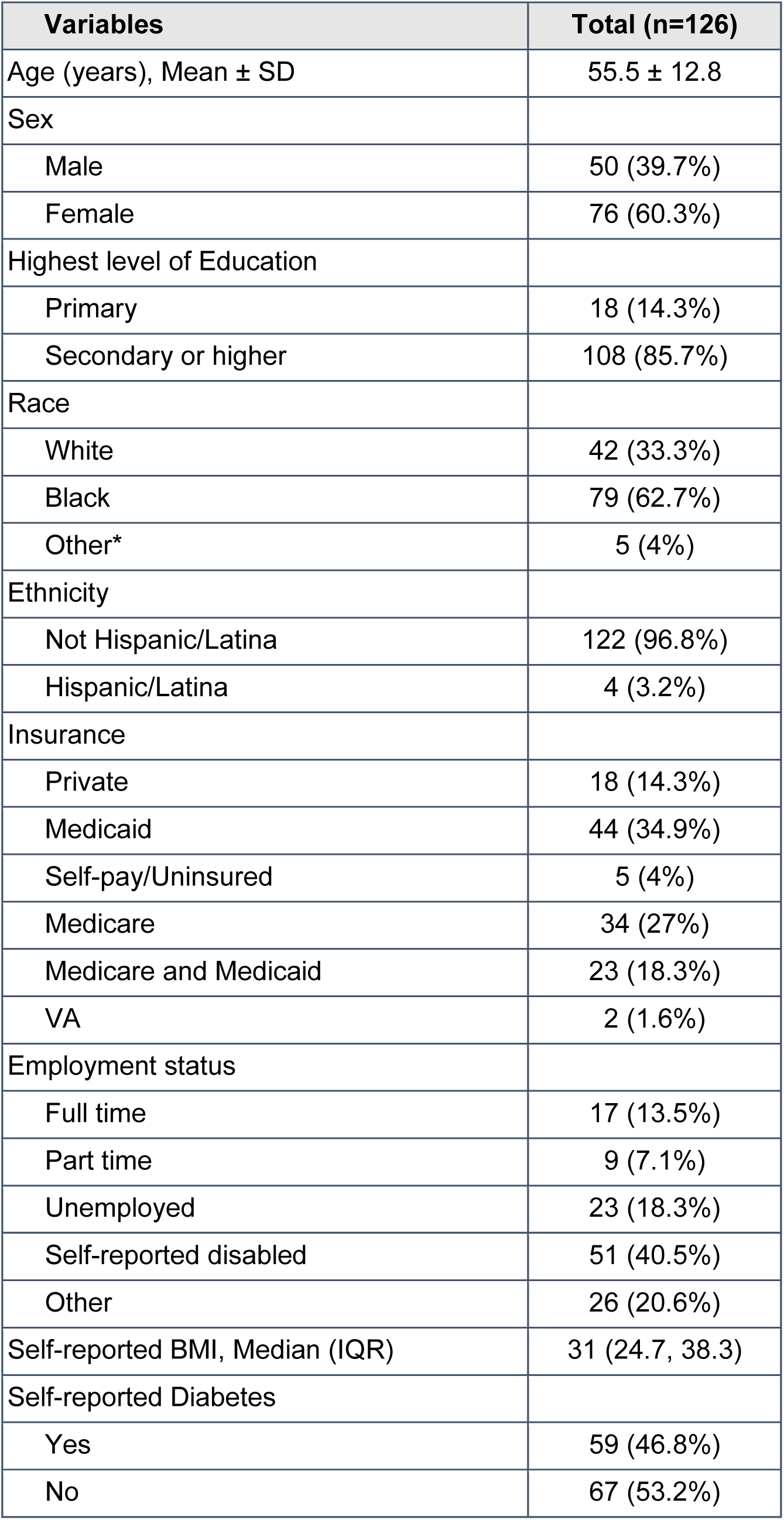
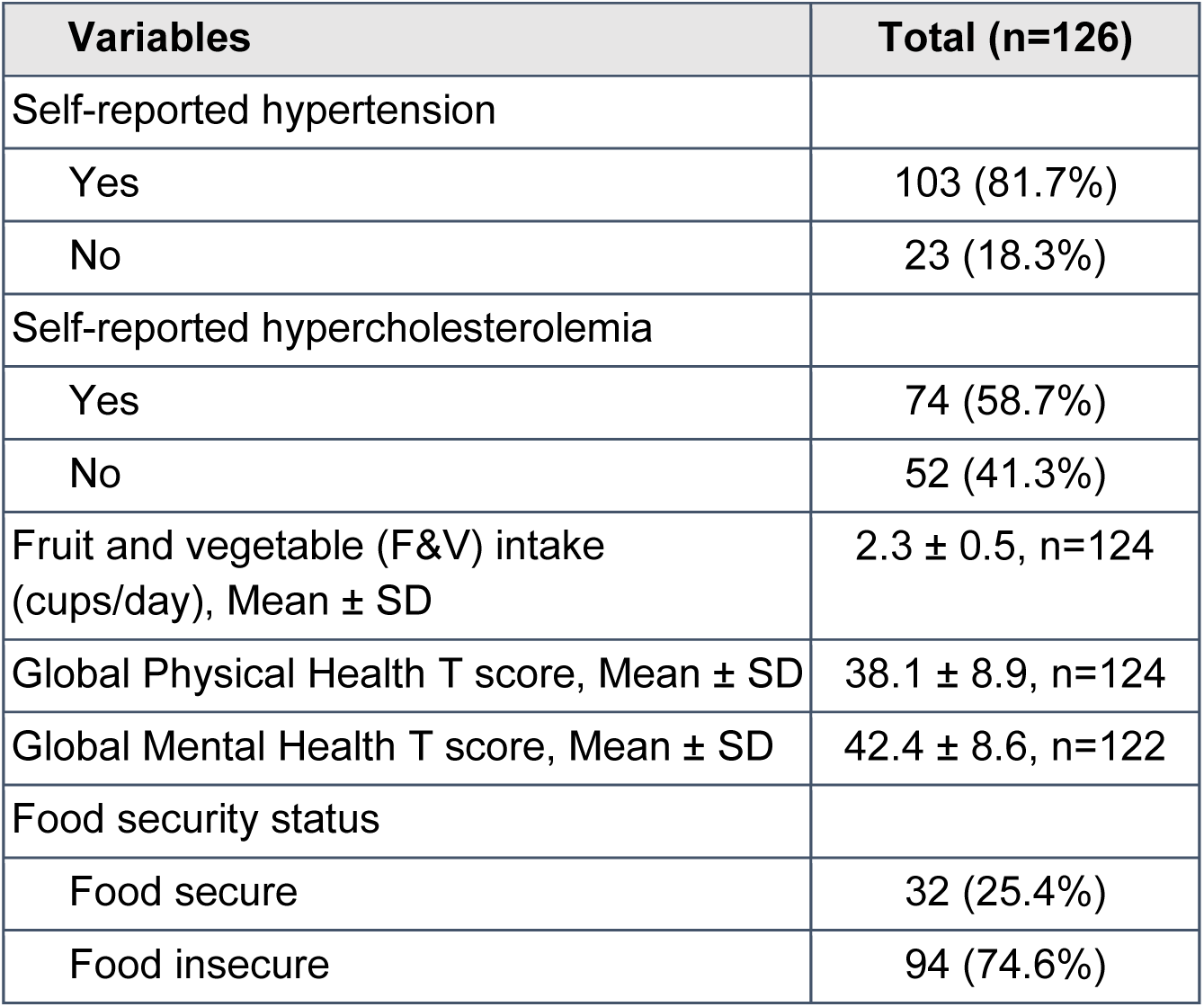
Baseline characteristics of NutriConnect participants.

| **Variables** | **Total (n=126)** |
| --- | --- |
| Age (years), Mean ± SD | 55.5 ± 12.8 |
| Sex |  |
| Male | 50 (39.7%) |
| Female | 76 (60.3%) |
| Highest level of Education |  |
| Primary | 18 (14.3%) |
| Secondary or higher | 108 (85.7%) |
| Race |  |
| White | 42 (33.3%) |
| Black | 79 (62.7%) |
| Other* | 5 (4%) |
| Ethnicity |  |
| Not Hispanic/Latina | 122 (96.8%) |
| Hispanic/Latina | 4 (3.2%) |
| Insurance |  |
| Private | 18 (14.3%) |
| Medicaid | 44 (34.9%) |
| Self-pay/Uninsured | 5 (4%) |
| Medicare | 34 (27%) |
| Medicare and Medicaid | 23 (18.3%) |
| VA | 2 (1.6%) |
| Employment status |  |
| Full time | 17 (13.5%) |
| Part time | 9 (7.1%) |
| Unemployed | 23 (18.3%) |
| Self-reported disabled | 51 (40.5%) |
| Other | 26 (20.6%) |
| Self-reported BMI, Median (IQR) | 31 (24.7, 38.3) |
| Self-reported Diabetes |  |
| Yes | 59 (46.8%) |
| No | 67 (53.2%) |

| Variables | Total (n=126) |
| --- | --- |
| Self-reported hypertension |  |
| Yes | 103 (81.7%) |
| No | 23 (18.3%) |
| Self-reported hypercholesterolemia |  |
| Yes | 74 (58.7%) |
| No | 52 (41.3%) |
| Fruit and vegetable (F&V) intake<br>(cups/day), Mean ± SD | 2.3 ± 0.5, n=124 |
| Global Physical Health T score, Mean ± SD | 38.1 ± 8.9, n=124 |
| Global Mental Health T score, Mean ± SD | 42.4 ± 8.6, n=122 |
| Food security status |  |
| Food secure | 32 (25.4%) |
| Food insecure | 94 (74.6%) |

### Primary Outcome

The primary outcome of the NutriConnect is the between group difference in change in F&V intake from baseline to the 6-month follow-up among individuals randomized to the Delivery and Credit arms. F&V consumption is measured in cups per day using a validated 10-item dietary assessment tool derived from the 26-item Dietary Screener Questionnaire (DSQ) developed by the National Cancer Institute.^17^ The study hypothesizes that participants in the NutriConnect Delivery will experience a greater increase in F&V intake compared to those in the NutriConnect Credit at 6 months.

### Secondary Outcomes

The secondary outcomes of the NutriConnect are the between group differences in F&V intake from baseline to the 6-month, changes in household food security, and self-reported health-related quality of life among individuals randomized to all three arms. Household food security is assessed using the 6-item USDA U.S. Household Food Security Survey (30-day version),^18^ with changes categorized as binary (food secure vs. food insecure). Self-reported health-related quality of life is measured using the Patient Reported Outcomes Information System (PROMIS) Global Health 5-point Likert scale,^19^ capturing participants’ perception of their physical and mental health at baseline and 3- and 6-month follow-up.

### Implementation Outcomes

In addition to primary and secondary measures, the study will evaluate implementation outcomes to assess the reach, implementation, cost, maintenance, and sustainability of NutriConnect using the PRISM/RE-AIM framework (**Table 2).**^20^ Implementation outcomes will be primarily descriptive and explore potential factors associate with implementation, scale-up, and sustainability. We will conduct key informant interviews with patients as well as hospital frontline, administrators, and community partner members. The study purposively will select a sample of participants (∼12 patients overall, with variability across by age, sex, race/ethnicity) from each of the NutriConnect active arms. Patient interviews will be conducted in conjunction with the 3-month follow-up visit. Interviews with hospital frontline (n=10), administrators (n=5) and Schnucks staff (n=4) will be conducted at the conclusion of intervention. Semi-structured interviews are guided by PRISM/RE-AIM (**Table 2**).^20^

**Table 2.** Schedule of trial follow-up and data collection.

|  | Screening<br>(Pre-consent) | Phone Contact<br>(Recruitment,<br>Baseline) | Month 3 | Month 6 |
| --- | --- | --- | --- | --- |
| EMR Review Eligibility | X |  |  |  |
| Informed Consent |  | X |  |  |
| Socio-Demographics |  | X |  |  |
| Outcome Evaluation |  |  |  |  |
| Randomization |  | X |  |  |
| Consumption of Fruit & Vegetables<br>(F&V) |  | X | X | X |
| Household Food Insecurity |  | X | X | X |
| Self-Reported Health Status |  | X | X | X |
| Control & Experimental Interventions<br>(Credit or Delivery)<br>Frequency: every two weeks; Duration of<br>intervention: 6 months |  | X |  | X |
| Interview on program perspectives<br>(sampled patients) |  |  | X |  |

**Table 3.** RE-AIM/PRISM Construct for Implementation Outcome Assessments.

| **Construct** | **Measure** |
| --- | --- |
| *RE-AIM* |  |
| Reach | Process data (e.g., number of potential patients contacted, enrolled) |
| Adoption | Hospital frontline staff interviews |
| Implementation Fidelity | Hospital frontline staff interviews; study team observation, team huddle |
| Implementation Cost | Exploratory outcome |
| Effectiveness | Effectiveness outcome 6-months post-intervention |
| Maintenance | Follow-up post intervention (planned, but not formal part of NutriConnect) |
| *PRISM* |  |
| Organizational perspective | Hospital frontline staff, administrator, and Schnucks staff interviews |
| Patient perspective | Patient interviews |
| External environment | Hospital frontline staff, administrator, and Schnucks staff interviews |
| Implementation and sustainability | Program Sustainability Assessment Tool (PSAT) |
| Infrastructure | Hospital frontline staff, administrator, and Schnucks staff interviews |
| Organizational characteristics | Hospital frontline staff, administrator, and Schnucks staff interviews |
| Patient characteristics | Patient baseline sociodemographics survey |

### Exploratory Outcomes

The study also explores changes in shopping habits and implementation costs. For all participants, grocery purchasing behavior will be analyzed using Schnucks Rewards shopping records, specifically tracking F&V purchases. To evaluate program feasibility, time-driven activity-based costing will be used to estimate the costs associated with NutriConnect implementation from a healthcare system perspective, incorporating structured activity logs and personnel time tracking. Research staff are conducting monthly reviews of project records and collect self-reported activity logs from hospital personnel and Schnucks personnel.^21^ Costs are tabulated for each activity related to implementation. Time spent per activity is tabulated for the personnel involved. The total activity cost will be calculated based on the number of personnel involved and estimated hourly wages base on national averages from the Bureau of Labor Statistics.^22^ Any expenses associated with implementation activities will also be included.

## Date Analysis

### Sample size and power

Power computations were based on the primary endpoint the difference between NutriConnect Delivery and Credit in the between group difference in mean change in F&V intake at 6 months. We hypothesize that the Delivery arm will have higher change in mean F&V intake compared to the Credit arm at 6-months. The power analysis was based on a two-sample t-test to evaluate the null hypothesis of no difference between Credit and Delivery arms (H0:μDelivery− μCredit = 0). For the Delivery arm, we anticipated a minimum additional increase of 0.5 cups per day in F&V intake compared to the Credit arm due to greater convenience and accessibility for the target population. This difference falls well within the range of similar food-prescription interventions. Our results indicated that after accounting for 20% attrition, we needed 80 patients per arm (n=240) to detect an effect size of 0.5 or more with 80% power at a two-sided alpha = 0.05. However, the study team was not able to achieve this sample size over the recruitment period due to greater than anticipated outreach challenges, including low response to initial post-discharge contact and participant concerns about randomization into the usual care group.

### Statistical Analysis

All analyses will be performed following a modified intention-to-treat approach, including all randomized participants who completed a baseline assessment and have at least one follow-up measure. In this analysis, participants remain in the groups to which they were assigned, regardless of adherence. The primary endpoint is the difference in change in mean daily F&V intake (cups/day) between the Delivery and Credit arms at six months. A linear mixed-effects model will be used to account for repeated measurements three months and six months. Fixed effects will include treatment group (Delivery, Credit, usual care), time (categorical), and baseline covariates (e.g., age, sex, baseline F&V intake), as well as a group-by-time interaction to estimate differential changes across groups over time. A random intercept for each participant will capture within-participant correlations. The main contrast of interest is Delivery versus Credit at six months, although each arm will also be compared separately and collectively against enhanced usual care as a secondary comparison. Adjusted mean differences and 95% confidence intervals will be estimated for each contrast. All hypothesis tests will be two-sided with an alpha of 0.05 with no adjustment for multiple testing. A p-value < 0.05 for the primary endpoint (i.e., Delivery vs. Credit on F&V intake at six months) will be considered a statistically significant difference.

For the secondary outcomes, self-reported health-related quality of life will be analyzed with the same linear mixed-effects approach as for the primary endpoint. Household food security will be analyzed using a modified Poisson with robust standard errors.^23^ All outcomes will be compared among treatment arms at both three and six months. Sub-group analyses stratified by age, race/ethnicity, education, baseline F&V intake, geographic proximity to Schnucks will be conducted to evaluate for potential heterogeneity in treatment effects. For each subgroup, we will include a treatment-by-subgroup interaction term in the primary model, using p < 0.10 to account for reduced statistical power in detecting heterogeneity. If we find evidence for effect modification, we will conduct additional exploratory stratified analyses to estimate and compare treatment effects within each subgroup.

### Baseline Characteristics

NutriConnect participants (*n*=126) had a mean age of 55.5 years (standard deviation [SD] = 12.8) and were predominantly female (60.3%) (**Table 1**). The majority had at least a secondary level of education (85.7%) and identified as Black (62.7%). Medicaid (34.9%) and Medicare (26.2%) were the most common forms of insurance coverage. Employment status varied, with 40.5% self-reporting as disabled. Full-time and part-time employment were less common, reported by 13.5% and 7.1% of participants, respectively. The median BMI was 31 (IQR: 24.7– 38.3), and nearly half (46.8%) reported having diabetes, 81.7% had hypertension, and 58.7% reported hypercholesterolemia. The average daily intake of fruits and vegetables was 2.3 cups (SD = 0.5). Notably, 74.6% of the sample reported experiencing food insecurity.

### Conclusion

In conclusion, the NutriConnect represents an important step toward addressing food insecurity and improving dietary intake among socioeconomically disadvantaged populations with diet-sensitive conditions by comparing two produce prescription strategies. By comparing the effect and implementation of the NutriConnect Credit and NutriConnect Delivery arms, this trial will provide valuable insights into strategies to improve fruit and vegetable consumption, food security, and self-reported health outcomes. Given the persistent disparities in food access and diet-related health conditions, findings from this research may inform future trials, programs, and policies to scale-up sustainable produce prescription programs.

## Data Availability

All data produced in the present study will be available upon reasonable request to the authors.

## References

1. Gondi KT, Larson J, Sifuentes A, et al. Health of the food environment is associated with heart failure mortality in the United States. Circ: Hear Fail. 2022;15(12):e009651. doi:10.1161/circheartfailure.122.009651

2. Clark AM, DesMeules M, Luo W, Duncan AS, Wielgosz A. Socioeconomic status and cardiovascular disease: risks and implications for care. Nat Rev Cardiol. 2009;6(11):712–722. doi:10.1038/nrcardio.2009.163

3. Leng B, Jin Y, Li G, Chen L, Jin N. Socioeconomic status and hypertension. J Hypertens. 2015;33(2):221–229. doi:10.1097/hjh.0000000000000428

4. Seligman HK, Laraia BA, Kushel MB. Food insecurity is associated with chronic disease among low-income NHANES participants 1, 2. J Nutr. 2010;140(2):304–310. doi:10.3945/jn.109.112573

5. Volpp KG, Berkowitz SA, Sharma SV, et al. Food is medicine: a presidential advisory from the American Heart Association. Circulation. 2023;148(18):1417–1439. doi:10.1161/cir.0000000000001182

6. Downer S, Berkowitz SA, Harlan TS, Olstad DL, Mozaffarian D. Food is medicine: actions to integrate food and nutrition into healthcare. BMJ. 2020;369:m2482. doi:10.1136/bmj.m2482

7. Hager K, Shi P, Li Z, et al. Evaluation of a produce prescription program for patients with diabetes: a longitudinal analysis of glycemic control. Diabetes Care. 2023;46(6):1169–1176. doi:10.2337/dc22-1645

8. Bhat S, Coyle DH, Trieu K, et al. Healthy food prescription programs and their impact on dietary behavior and cardiometabolic risk factors: a systematic review and meta-analysis. Adv Nutr. 2021;12(5):nmab039-. doi:10.1093/advances/nmab039

9. Auvinen A, Simock M, Moran A. Integrating produce prescriptions into the healthcare system: perspectives from key stakeholders. Int J Environ Res Public Heal. 2022;19(17):11010. doi:10.3390/ijerph191711010

10. DePuccio MJ, Garner JA, Hefner JL, Coovert N, Clark A, Walker DM. Multi-stakeholder perspectives on the implementation of a clinic-based food referral program for patients with chronic conditions: a qualitative examination. Transl Behav Med. 2022;12(9):927–934. doi:10.1093/tbm/ibac027

11. Houghtaling B, Greene M, Parab KV, Singleton CR. Improving fruit and vegetable accessibility, purchasing, and consumption to advance nutrition security and health equity in the United States. Int J Environ Res Public Heal. 2022;19(18):11220. doi:10.3390/ijerph191811220

12. Gao Y, Yang A, Zurbau A, Gucciardi E. The effect of food is medicine interventions on diabetes-related health outcomes among low-income and food-insecure individuals: A Systematic Review and Meta-analysis. Canadian Journal of Diabetes. 2023;47:143–152. doi:10.1016/j.jcjd.2022.11.001

13. Hager K, Du M, Li Z, et al. Impact of produce prescriptions on diet, food security, and cardiometabolic health outcomes: a multisite evaluation of 9 produce prescription programs in the United States. Circ: Cardiovasc Qual Outcomes. 2023;16(9):e009520. doi:10.1161/circoutcomes.122.009520

14. MOST Policy Initiative. Tax Credits for Grocery Stores in Food Deserts. Accessed March 6, 2025. https://mostpolicyinitiative.org/wp-content/uploads/2022/01/Tax-Credits-Grocery-Stores-V2.pdf

15. Food Bank. Hunger Facts & Stats. https://stlfoodbank.org/hunger-facts/

16. Hager ER, Quigg AM, Black MM, et al. Development and validity of a 2-item screen to identify families at risk for food insecurity. Pediatrics. 2010;126(1):e26–e32. doi:10.1542/peds.2009-3146

17. National Cancer Institute. Dietary Screener Questionnaires (DSQ). 2021. Accessed March 6, 2025. https://epi.grants.cancer.gov/nhanes/dietscreen/questionnaires.html

18. Nord M. A 30-day food security scale for current population survey food security supplement data. Food Assistance & Nutrition Research Program; 2002. Accessed March 6, 2025. https://ers.usda.gov/sites/default/files/_laserfiche/publications/43192/31177_efan02015_002.pdf?v=13323

19. Hays RD, Bjorner JB, Revicki DA, Spritzer KL, Cella D. Development of physical and mental health summary scores from the patient-reported outcomes measurement information system (PROMIS) global items. Qual Life Res. 2009;18(7):873–880. doi:10.1007/s11136-009-9496-9

20. Feldstein AC, Glasgow RE. A practical, robust implementation and sustainability model (PRISM) for integrating research findings into practice. Jt Comm J Qual Patient Saf. 2008;34(4):228–243. doi:10.1016/s1553-7250(08)34030-6

21. Kaplan RS, Anderson SR. The innovation of time-driven activity-based costing. Cost Management. 2007;(21):5–15.

22. U.S. Bureau of Labor Statistics. Occupational Employment and Wage Statistics. Accessed May 6, 2025. https://www.bls.gov/oes/

23. Zou G. A modified poisson regression approach to prospective studies with binary data. Am J Epidemiology. 2004;159(7):702–706. doi:10.1093/aje/kwh090

